# Data Quality Assurance Tool for the Acute to Chronic Pain Signatures Study (A2CPS): An Interactive R Shiny Application

**DOI:** 10.64898/2026.01.07.26343620

**Authors:** Briha Ansari, Patrick Sadil, James Ford, Giovanni Berardi, Margaret Taub, Ari Kahn, Joshua Urrutia, Andre Hackman, Adi Gherman, Martin A. Lindquist, The Acute to Chronic Pain Signatures Consortium

## Abstract

**Background/Aims:** Clinical trials and observational studies support the synthesis and development of clinical guidelines, highlighting the need for strong data quality assurance measures. The Acute to Chronic Pain Signatures (A2CPS) program is a large-scale, multi-site observational study investigating chronic post-surgical pain and opioid dependence. Its primary goal is to identify biomarkers predictive of progression from acute to chronic pain following knee arthroplasty or thoracic surgery. The A2CPS sites collect data across various domains, including brain magnetic resonance imaging, electronic health records, psychosocial measures, multi-omics, Quantitative Sensory testing, and functional testing.

While A2CPS is an observational study, its aims, design, and methodology closely align with clinical trial practices. This includes interdisciplinary collaboration, standardized protocols, defined eligibility criteria, and oversight by a Data and Safety Monitoring Committee.

In multifaceted studies like A2CPS, high-quality data are paramount to ensure the accuracy of predictive biomarkers. To improve quality assurance, we developed the A2CPS Data Monitoring Web Application (Web App), an interactive R Shiny web app with real-time data monitoring capabilities. Here, we describe the functionality and utility of the A2CPS Data Monitoring Web App in streamlining quality assurance for the A2CPS study.

**Methods:** The Web App is a secure R Shiny web application accessible to authorized A2CPS Data Integration and Resource Center (DIRC) members. It retrieves and preprocesses data from REDCap, which is then fed into the R Shiny framework. The user interface has a navigation bar and six subpanels, providing easy access to the app’s modules and enabling users to switch seamlessly among subpanels. Each subpanel addresses a specific use case and has the functionality to generate downloadable error reports for individual sites, making it easy to share quality documents and communicate with data collection sites. The DIRC uses these reports to identify errors, coordinate remediation, and facilitate targeted training for research personnel.

**Results:** Regular use of the Web App, coupled with engagement with the training team, resulted in an overall reduction of 50% in data quality errors over one year in case report form data (i.e., in-person visit data). The decline in errors was consistent across all sites despite steady enrollment rates, indicating that real-time data monitoring enables focused feedback, mitigates recurring errors, and streamlines data quality assurance.

**Conclusion:** The A2CPS Data Monitoring Web App plays a key role in A2CPS data quality assurance. This robust open-source solution reduces data entry errors and provides targeted feedback and training to the data collection sites. Our results demonstrate the potential for using open-source computational frameworks for data monitoring and quality assurance purposes in both clinical trials and observational studies.

## Introduction

Clinical trials and observational studies support the synthesis and development of clinical guidelines, highlighting the need for robust data quality assurance measures^1^. Indeed, multiple studies have shown that data quality determines the reliability, accuracy, and generalizability of research findings. Although data quality assurance can pose a significant economic burden (e.g., personnel hours), the benefits outweigh the costs in research practice^2^.

These considerations are particularly true for studies such as the Acute to Chronic Pain Signatures (A2CPS) study, a large-scale, multisite observational study investigating chronic post-surgical pain and opioid dependence in 2,800 post-surgical patients^3^. The main goal of A2CPS is to identify biomarkers predictive of progression from acute to chronic pain in patients following knee arthroplasty (TKA) or thoracic surgery. The A2CPS program includes two Multisite Clinical Centers (MCC1 and MCC2), each consisting of multiple sites that recruit and enroll both surgical cohorts and collect data across various domains, including psychosocial measures, brain magnetic resonance imaging (MRI), electronic health records (EHR), multi-omics, Quantitative Sensory Testing (QST), and functional testing. Psychosocial data is remotely collected as Patient Reported Outcomes (PROs), while case report form (CRF) data from brain MRIs, biospecimen collection (multi-omics), functional testing, and QST are collected in person in a dedicated clinical setting by research staff. Please refer to the A2CPS Study protocol paper for more information^3^.

Although A2CPS is an observational study, it shares many similarities to a clinical trial. Like multi-center clinical trials, A2CPS is an interdisciplinary initiative bringing together experts from various fields to guide decision-making and unravel the mechanisms driving chronic pain — a complex medical condition. A2CPS follows standardized protocols for data collection across all its sites, has defined eligibility criteria for study participants, and has a data and safety monitoring committee, all of which are standard practice in clinical trials. The dissemination of findings is an important aspect of clinical trials; similarly, the A2CPS study data will be shared with the broader scientific community to foster reproducibility and future research^4^.

In multifaceted studies like A2CPS, high-quality data is paramount to ensure accurate, generalizable predictive modeling across diverse populations. Day-to-day operations require real-time data monitoring capabilities to help recognize errors as they evolve. Early detection of errors enables timely communication with the sites and aids in designing targeted interventions (e.g., changes to PROs and CRFs, or additional training) to offset the possibility of future errors or protocol drift, and to improve data quality.

To support these goals, we developed the A2CPS Data Monitoring Web App with real-time data monitoring capabilities. The app was built with the R Shiny framework, which leverages packages in the R ecosystem to facilitate building customizable, user-friendly Web Apps^5^. The A2CPS Data Monitoring Web App integrates a back end for data retrieval, processing, and report generation for each site for use by the A2CPS Data Integration and Resource Center (DIRC) via a user-friendly interface with navigation menus and interactive elements for data visualization. In this paper, we describe the functionality of the A2CPS Data Monitoring Web App and its utility in streamlining quality assurance for the A2CPS study.

## Methods

### Data Management and Security

A2CPS employs a multiprong approach to data security and storage. A2CPS uses Research Electronic Data Capture (REDCap), a secure web-based application for electronic data collection, storage, and dissemination^6^. REDCap provides a common interface that allows data entry from multiple sites.

The DIRC, based at Johns Hopkins University, works with the Texas Advanced Computing Center (TACC) at the University of Texas to store and analyze the data. A unique study identification number is assigned to each subject to obfuscate personally identifiable health information (PHI).

REDCap is hosted on a centrally managed local server at TACC. Data can be programmatically exported from REDCap using REDCap application programming interface (API) calls for use by various statistical software platforms such as Excel, SAS, and R^6^.

The A2CPS Data Monitoring Web App runs as a desktop application accessed by authorized DIRC users via the TACC virtual private network (VPN), which protects sensitive information from unauthorized access in transit. The Web App retrieves data from REDCap using an API call and a robust, token-based authentication system: to initialize the app, users enter a personal API token (assigned to authorized users only). The API also attaches metadata to keys and data to prevent unauthorized data manipulation^7^.

### Ethics Statement

This study was approved by the University of Iowa Institutional Review Board, serving as the central IRB. All patients/participants provided their written informed consent prior to study participation

### The A2CPS Data Monitoring Web Application Description

The A2CPS Data Monitoring Web App retrieves A2CPS (CRF/PRO) data from REDCap via an API call, retrieved data are then processed and validated using a series of quality checks defined by the A2CPS data curation team. These checks include identifying missingness in PRO data and data entry errors in the CRFs. On average, approximately 20 quality checks are performed per CRF per site. Table 1 includes examples of the checks for each CRF.

**Table 1.** Examples of data quality checks by case report form and patient-reported outcomes.

| CRF(s)/PRO(s) | Data Quality Checks |
| --- | --- |
| Patient Reported Outcomes (PROs) | Check for missing data. |
| Quantitative Sensory Testing (QST) | Check for missing or mismatched Pressure Pain Threshold (PPT) ratings for the remote site; check if the Pressure Pain Threshold (PPT) was performed at the remote site but the information was entered for the index site. |
| Imaging | In subjects with a resting-state scan performed, check if there are missing values for surgical site pain or if there is a mismatch between entries for surgical site pain; after the first cuff fMRI scan (personalized cuff pressure), check if there are missing values or mismatches in cuff pain ratings during the mid-scan. |
| Blood Sample Collection and Processing | Check if there is missing information on vaccination; check for discrepancies in blood draw time, centrifuge time, and freezer time. |
| Functional Testing | Check for discrepancies or missing values between the first and second initial pain ratings; check if the reason for the test not being completed was not specified. |

The data generated from quality checks are then organized into structured tables, where columns represent variables, rows represent observations, and each cell contains a single value^8^. These tables are then fed into the web app’s user interface (UI) and server component for data visualization at the front end. Due to the intensive data preprocessing required at the Web App’s back end, users should anticipate significant RAM usage and extended processing times when running the app locally.

The app’s front end has a main panel with six subpanels and a user-friendly navigation bar at the top that allows users to navigate between the sub-panels seamlessly. Each subpanel represents a specific use case and displays output customizable by the dropdown menu in the sidebars. The script corresponding to each subpanel expects a table that contains columns with input parameters for the sidebar to generate plots in the output field. Incorrect formatting of the table will impair the Web App’s functionality. This can be offset by organizing data into tables containing variables with the input parameters for the UI. Each subpanel has the functionality to export plots and site-level error reports, making it easy to share quality documents with collaborators.

As the study is ongoing and data have not yet been fully released to the public, we have used images from the Web App based on simulated data for this paper. The following sections describe the layout and functionality of each subpanel in more detail.

### Reproducible Example of the A2CPS Data Monitoring Web App availability

A link to the reproducible example, along with the source code for the A2CPS Data Monitoring Web App using simulated data is provided in the Data and Code Availability section.

### Subpanel 1: Data Completion and Error Overview

This subpanel summarizes data completion by visit type and site for each CRF/PRO and provides an overview of CRF/PROs completed without errors (Fig. 1A). A four-quadrant display reacts to the user input in the sidebar. The sidebar includes three dropdown menus that allow concurrent selection of multiple visits and sites for each CRF/PRO, making comparisons across the various time points and sites more efficient. The four-quadrant main panel employs bar plots to show: (1) percentage of CRFs/PROs completed without errors; (2) percentage of CRFs/PROs completed with errors; (3) percentage of incomplete CRFs/PROs; and (4) the percentage of CRFs/PROs with unknown completion status.

**Figure 1.**
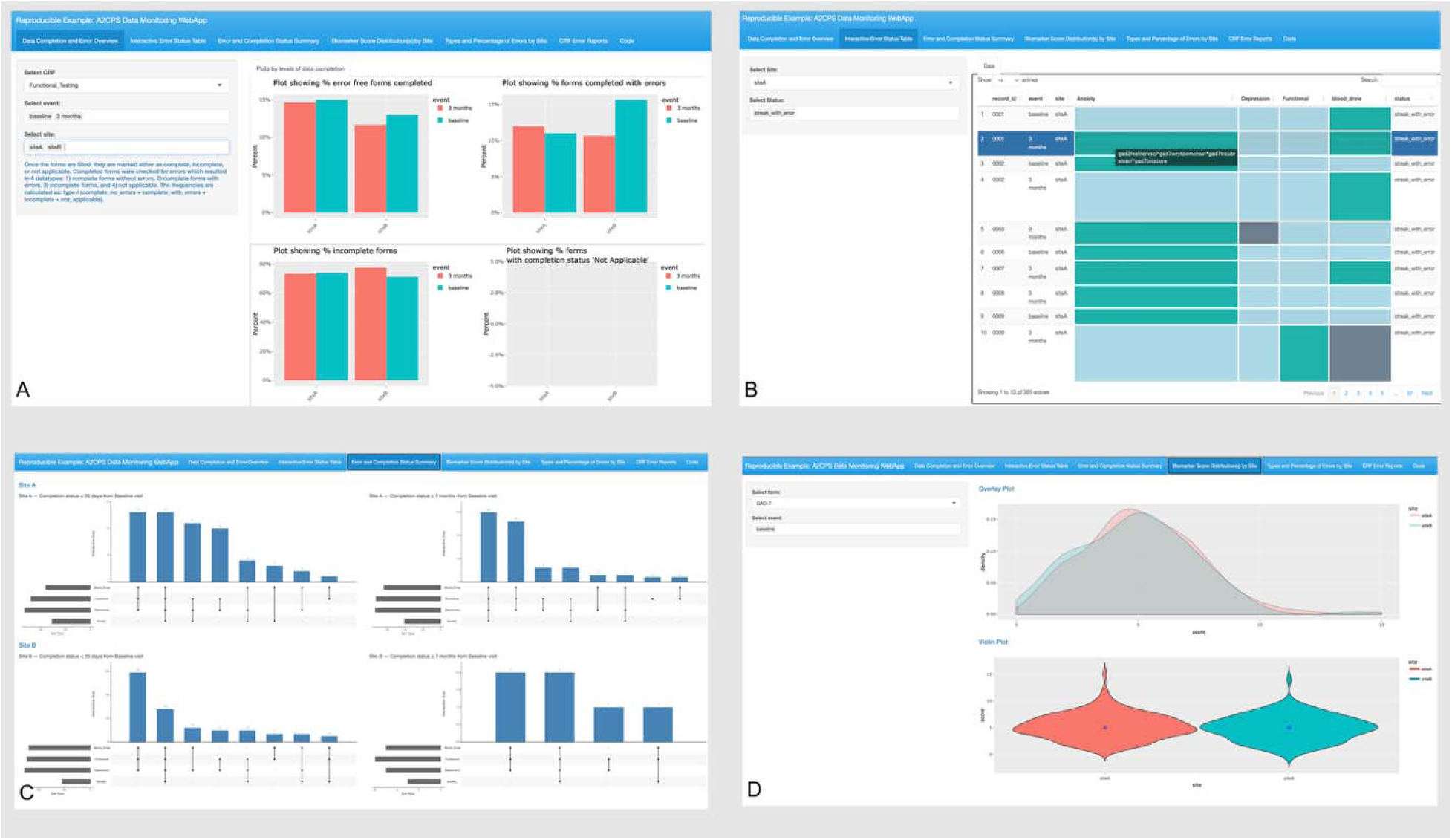
First four subpanels of the A2CPS Data Monitoring Web Application. (A) Subpanel 1: Four-quadrant display showing the percentage of forms completed without errors, completed with errors, incomplete forms, and forms with unknown completion status, stratified by site and visit type. (B) Subpanel 2: Interactive table highlighting subject-level errors across different case report forms and patient-reported outcomes by visit type. (C) Subpanel 3: UpSet plots visualizing the number of subjects who completed various combinations of case report forms and patient-reported outcomes without errors. (D) Subpanel 4: Biomarker score distributions for each case report form and patient-reported outcome by visit type and surgical cohort.

### Subpanel 2: Interactive Error Status Table

This subpanel uses an interactive table to highlight subject level errors across different CRF/PROs by visit type (Fig. 1B). The purpose of this subpanel is to provide a granular view of subject level errors. The interactive error status table in the output field reacts to the user input in the sidebar. The sidebar comprises two dropdown menus, allowing simultaneous selection of site and of the following statuses: (1) subjects with all primary biomarkers available across time with no errors; (2) subjects with all primary biomarkers available across time with errors (streak with errors); and (3) subjects with incomplete biomarker data (incomplete streak).

The main panel displays an interactive table where rows represent subject IDs and columns correspond to specific CRF/PROs associated with each subject. Each cell in the table is color-coded, where blue indicates error-free data, green indicates data with errors, and yellow indicates data not collected by design. The table uses tooltips (hover-over text) to provide additional information about errors in green cells.

### Subpanel 3: Error and Completion Status Summary

This subpanel uses an UpSet plot^9^ to visualize the number of subjects who have completed various combinations of CRF/PROs without errors, highlighting the number of subjects with complete biomarker data (Fig. 1C). Each vertical bar represents a specific combination of completed CRF/PROs. For example, a bar might show subjects who completed their neuroimaging and functional testing but not GAD-7 (Questionnaire for anxiety) and PHQ-9 (Questionnaire for depression). The horizontal bar length indicates the total number of subjects who have completed each CRF/PRO. There is no sidebar for user input since the UpSet plots are static. The main panel shows UpSet Plots for each site arranged according to the time elapsed from baseline: the left side shows subjects at less than 35 days after the baseline visit and the right-side shows those at more than 7 months after the baseline visit. The former time point is chosen with the expectation that all pre-surgery surveys and immediate post-surgery pain trajectories (completed daily for 28 days) should be complete, allowing for an inventory of data completion. The latter time point similarly corresponds to the end of the window for completing primary endpoint surveys.

### Subpanel 4: Biomarker Scores Distributions

This subpanel presents overlay density and violin plots to illustrate the biomarker score distributions for each CRF/PRO by visit and surgical cohort (Fig. 1D). The subpanel aims to identify outliers in the scores and compare biomarker score distributions between the TKA and thoracic cohorts. The sidebar consists of two dropdown menus: one allows the user to select the CRF/PRO, while the second allows the user to choose the visit type.

### Subpanel 5: Types and Percentage of Errors by Site

MCC1 sites employ the REDCap survey delivery system, while MCC2 sites use MyDataHelps, an app-based survey delivery system to collect PROs which are uploaded to REDCap. This subpanel displays bar plots to show percentage of PRO specific error types across the different sites (Fig. 2A). The purpose of this subpanel is to identify if missingness in certain PROs depends on the instrument of survey delivery. The plots react to user selection of the PRO questionnaire type in the sidebar.

**Figure 2.**
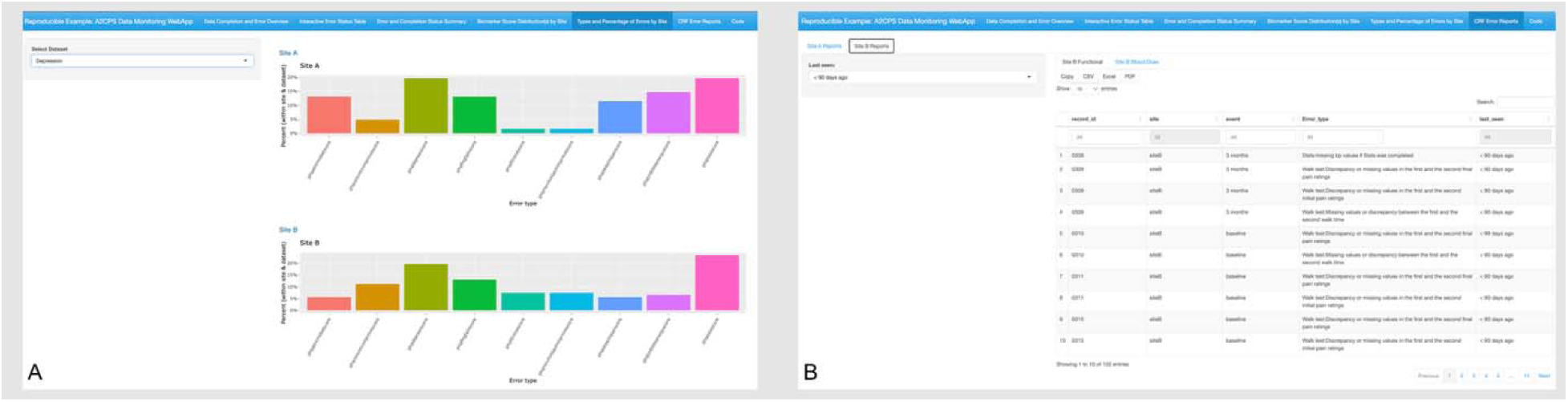
Last two subpanels of the A2CPS Data Monitoring Web Application. (A) Subpanel 5: Bar plots showing patient-reported outcome–specific error types across sites, used to assess whether missingness differs by survey delivery instrument. (B) Subpanel 6: Hierarchically structured panels displaying site-specific error reports with flagged subject identifiers for each case report form.

### Subpanel 6: CRF Error Reports

This panel has a hierarchical structure (Fig. 2B), with four distinct subpanels dedicated to each MCC and surgical cohort: MCC1 Knee arthroplasty, MCC1 thoracic surgery, MCC2 Knee arthroplasty, and MCC2 thoracic surgery. Within each subpanel, there are four nested CRF subpanels: neuroimaging, QST, functional testing, and biospecimen collection. The back end of the app checks CRF data for various errors and flags IDs with errors. The user can toggle between different CRFs, and each nested CRF subpanel will show an error report with flagged IDs and their error data. The sidebar comprises a dropdown menu with three separate time frames: < 90 days ago, 90 -180 days ago, and > 180 days ago. The error reports react to user selection and display errors within the specified time frame. This helps access the most recent errors while archiving older ones. The reports can be downloaded in PDF, Excel, or CSV format, and the DIRC uses these reports to communicate with the MCC sites to resolve any flagged data. The example app shows simulated data from two sites.

### Data Quality Assurance Process

The data collected from the MCC sites are entered into REDCap by trained research staff. REDCap is hosted on a centrally managed local server at TACC. An R script imports data from REDCap using an API call. The data are then preprocessed to generate error reports for each site. Processed data automatically feeds into the R Shiny app workflow, displaying the error reports and other data visuals through the R Shiny user interface for the A2CPS Data Monitoring Web App. The application does not require programming experience. Fig. 3 illustrates the overall quality assurance process and workflow.

**Figure 3.**
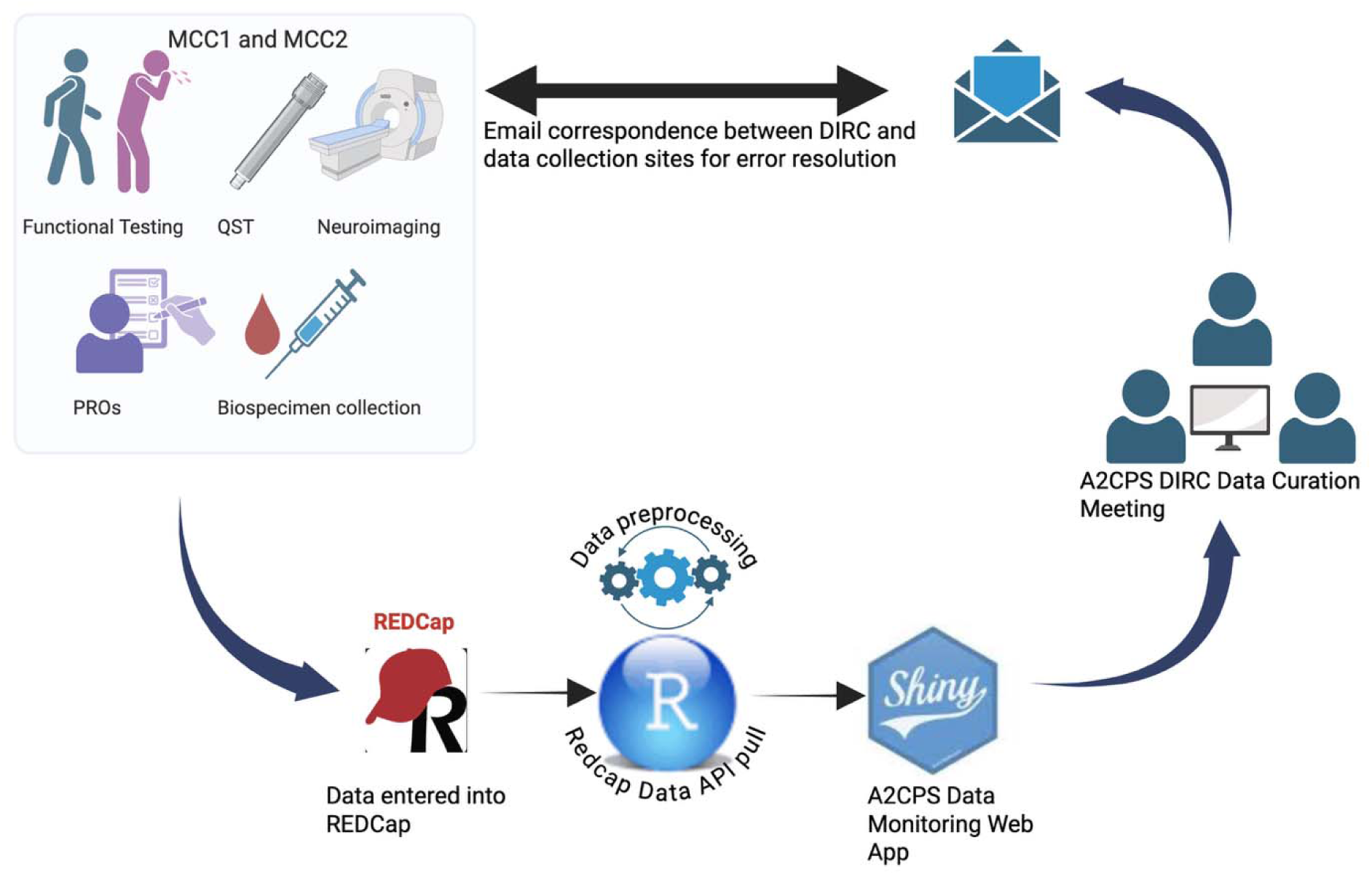
Data quality assurance process for the A2CPS Data Monitoring Web Application. This schematic illustrates the end-to-end workflow for data ingestion, quality checking, visualization, and site-level feedback used by the Data Integration and Resource Center. Created with BioRender.com. Ansari, B. (2026) https://BioRender.com/i32t673

DIRC members, including the principal investigator (PI), Martin Lindquist, meet biweekly for data curation meetings. During these meetings, the team uses the Web App to identify errors by CRF and site, brainstorm additional quality assurance checks, and address any recurring errors. The PI reviews and validates the errors in REDCap to determine if they can be resolved, maintains a log of the errors, and coordinates with the MCC sites to facilitate a resolution.

## Results

Routine use of the A2CPS Data Monitoring Web App with biweekly data curation meetings and followup interactions with the study’s training team has resulted in a 50% overall reduction in errors over one year, after accounting for variability in data quality errors across time and CRF(s). When aggregated across all CRF(s), the initial raw number of errors was 40 per month, which decreased to 6 per month at the most recent time point (with a similar enrollment rate), indicating an 85% reduction in data quality errors over time. The pooled estimate of 50% reduction was consistent with domain-specific models, although somewhat more conservative than the observed raw decrease from 40 to 6 errors (–85.0%). Within domains, Imaging and Blood Sample Collection and Processing showed the steepest model-estimated declines of 66.1% and 42.7%, respectively. Quantitative Sensory Testing errors decreased by 42.0%, while Functional Testing showed no consistent decline (–6.3%) (Table 2 and Fig. 4A and Fig. 4B).

**Figure 4.**
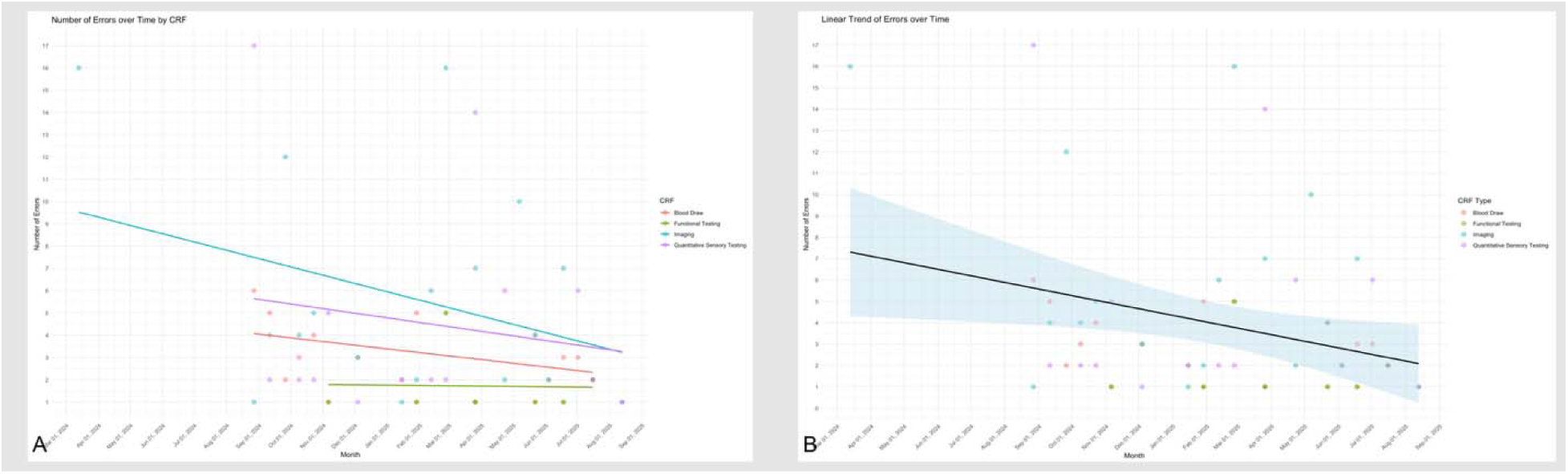
Reduction in data quality errors over time across case report forms. This figure summarizes changes in data quality errors following implementation of the A2CPS Data Monitoring Web Application. (A) Observed monthly error counts stratified by case report form, showing raw reductions in errors over time, and model-estimated percent reduction in data quality errors by domain, accounting for variability across time and case report forms. (B) Model-estimated overall percent reduction in data quality errors.

**Table 2.** Model-estimated reduction in data quality errors by domain.

| CRF | First date | First count | Last date | Last count | Observed % change | Model estimate % change |
| --- | --- | --- | --- | --- | --- | --- |
| Blood Sample Collection and Processing | 2024-08-27 | 6 | 2025-07-16 | 2 | -66.7% | -42.7% |
| Functional Testing | 2024-11-06 | 1 | 2025-07-16 | 2 | 100.0% | -6.3% |
| Imaging | 2024-03-13 | 16 | 2025-08-13 | 1 | -93.8% | -66.1% |
| Quantitative Sensory Testing | 2024-08-27 | 17 | 2025-08-13 | 1 | -94.1% | -42.0% |
| Total |  | 40 |  | 6 | -85.0% | -50.0% |

Observed % change reflects the raw difference between first and last count; model estimated % change was derived from domain specific models accounting for variability over time.

Subpanel 5 assesses whether missingness in PROs varies between the two survey delivery instruments, REDCap and MyDataHelps. To date, no difference has been observed.

The results demonstrate the pivotal role of ongoing feedback and training and suggest that implementing real-time data monitoring helps provide focused feedback, streamline the process of data quality assurance, and mitigate protocol drift and future errors. Furthermore, by automating error checks in REDCap, the Web App transforms a time prohibitive manual review of errors into a streamlined process that can be completed in under 30 minutes at biweekly intervals.

## Discussion

The success of the A2CPS Data Monitoring Web App underscores the significance of real-time monitoring to promote timely communication and targeted training of the research personnel at the data collection sites. Early detection of errors fosters constructive feedback and focused remediation, subsequently decreasing the error rate across sites.

Real-time data monitoring for quality assurance of clinical trials and observational studies helps offset human data quality errors and address unforseen or newly emerging errors, a common challenge in data management. Automated quality assurance systems have significantly enhanced the efficiency of data monitoring across various clinical domains. For example, Sundell et al. developed a real-time monitoring system for mammogram image quality, which improved diagnostic accuracy, clinical decision-making, and patient outcomes^10^. Similarly, ShinyLUTS applies R Shiny framework for structured data management and analysis in patients with lower urinary tract symptoms (LUTS)^11^.

To our knowledge, the A2CPS Data Monitoring Web App is one of the first data monitoring web applications developed using an open-source programming language for a longitudinal study. This paper focuses on real-time data monitoring and a user-friendly interface to aid the data curation team in recognizing error patterns. The Web App uses an API call to retrieve REDCap data and can be run as a desktop application by authorized users only. It uses a comprehensive library of R packages ^9, 12-28^ for data processing and generation of downloadable error reports. This allows the DIRC to consolidate site-specific error reports instead of reviewing REDCap data for errors, which is time consuming and costly.

Open-source software-based applications are a cost-effective option, but as with any software they come with challenges. Security and reliability will depend on organizational practices (e.g., access control and patching). For A2CPS, we guard against this by restricting access to DIRC users. Web Apps in particular can be limited in their ability to integrate into clinical database management platforms (e.g., REDCap and CASTOR EDC). Moreover, many software packages are not backed with a warranty, packages will vary in how and when they are updated. While updates typically improve functionality, they can result in unforeseen errors and bugs, requiring continuous cycles of upkeep, monitoring, and version control by the developers which can be time prohibitive^29^. Overall, based on the feedback of the A2CPS data curation team, implementing the A2CPS Data Monitoring Web App has improved the quality control and assurance of this large longitudinal study. This Web App leverages an open-source user friendly, R Shiny-based, computational framework that can be adapted by newer data monitoring and quality assurance applications in clinical trials and observational studies.

## Conclusion

In conclusion, the A2CPS Data Monitoring Web App plays a key role in A2CPS data quality assurance and training of research personnel. Since its introduction, data quality errors have decreased by 50%, highlighting the effectiveness of the Web App and the accompanying data curation process. Based on these findings, the A2CPS data monitoring Web App in combination with focused feedback and training, serves as a robust solution for reducing data entry errors and streamlining quality assurance workflows. The finding also suggests the potential for wider applicability of automated workflows in clinical research operations. In the future, we expect to improve this tool’s functionality and add more features to optimize the A2CPS data quality assurance.

## Data and Code Availability

A reproducible example, along with the source code for the A2CPS Data Monitoring Web App using simulated data is accessible at https://jhubiostatistics.shinyapps.io/a2cps-qc-app/.

## Acknowledgments

No AI text generation tool was used to help write this manuscript. However, Grammarly® basic features were used to check for spelling mistakes, typos, and grammatical errors. ChatGPT-5.0 was used to generate simulated data for the reproducible example of the A2CPS Data Monitoring Web App,

